# Evaluation of Knowledge, Practices, Attitude and Anxiety of Pakistan’s Nurses towards COVID-19 during the Current Outbreak in Pakistan

**DOI:** 10.1101/2020.06.05.20123703

**Authors:** SS Alwani, MM Majeed, MZ Hirwani, S Rauf, SM Saad, H Shah, F Hamirani

## Abstract

Since the emergence of the novel corona virus, the front line soldiers during this pandemic are the healthcare professionals because of their direct association with COVID19 patients. In the management of such patients, nurses play a significant role through proper care and preventive measures. Due to its contagious nature, fatality and no proper medicine, it is a risk to the health and life of nurses and has an impact on their psychological health.

In the current study we assessed the knowledge, attitude, practices and anxiety levels of nurses who are directly involved in the management of COVID-19 patients.

It was an online questionnaire based cross sectional survey targeting only those nurses involved in the management of COVID-19 patients from different hospitals of Karachi, Pakistan. SPSS 21 was used for data analysis. Descriptive analysis, Chi Square and t-tests were applied. P value < 0.05 was considered significant.

Data of 78 nurses was analyzed. We observed that nurses possess good knowledge about COVID-19, its sources, symptoms and routes of transmission of the Virus etc. The knowledge mean score was calculated 14.67 *±* 3.36. Health department /Hospital and social media are the main sources of information regarding COVID-19. We investigated that 92.3% of the nurses had mild to very severe anxiety and anxiety levels are significantly higher among females (P< 0.05).

We concluded that the nurses performing their duties with COVID-19 positive patients have good knowledge and attitude. But their anxiety levels are high. Psychological interventions along with training should be given.

## Introduction

The latest member of Corona Virus family is COVID-19 and before this two more members of the same family were the Severe acute respiratory syndrome coronavirus (SARSCoV) and Middle East respiratory syndrome coronavirus (MERSCoV) (1). The novel Corona virus (COVID-19) which was identified in December 2019 has been declared a pandemic on 11^th^ March 2020 as the virus was reported to be highly contagious and deadly (2). Based on the current data the total number of people infected and confirmed by the lab are more than 6 million and over 300000 have died while the figure is raising exponentially (3). In Pakistan, the first case of the virus was reported on 26^th^ Feb 2020, a university student travelled from Iran, which was the epi center of this virus after China (4). By this time (31^st^ May 2020) Pakistan has crossed the 70000 mark of reported cases and with more than 1400 deaths so far(5).

To control the spread, Centers for Disease Control and Prevention (CDC) has recommended a guideline and a protocol to follow(6). Mainly this virus spreads via aerosols, direct contact with the infected person or indirectly by touching surface that has virus on it. Fever, sore throat, dry cough and shortness of breath are the main symptoms (7). Up till now, there is no specific treatment other than supportive treatment and its morbidity rates are less than 5% (8, 9). The nature of this virus, is rapidly changing and new information regarding COVID-19 is pouring on a daily basis. So updated knowledge and training is very necessary in order to tackle this deadly virus(7).

Healthcare professionals (HCPs) are the front line soldiers in the outbreak of any disease and are more susceptible to be infected because of their direct and close interaction with the diseased individuals (10, 11). They may not only be infected but in severe cases it may lead to the death of the HCPs or a similar scenario may happen with their family members or closed ones (12). In the current pandemic, thousands of healthcare professionals and healthcare workers were infected in China, Spain, Italy, France, Turkey and other parts of the world(13–15). International council of Nursing (ICN) stated that over 90000 healthcare workers have been infected with COVID-19 and only the death toll of nurses is estimated to be 360 (16, 17). In Pakistan, hundreds of HCPs are reported COVID-19 positive along with many deaths (18, 19). Moreover, a suicide case of 27 years old doctor has also been reported in the turmoil of COVID 19 due to anxiety (20).

The current situation is a challenge for HCPs to cope up with anxiety, stress, and depression, not only for their health and life but for the safety of family members. In such conditions, it is not uncommon for the front line workers that includes medical doctors, dentists, nurses and other health care professions to develop psychological symptoms, mental and other health related problems (21). Health care workers involved with the testing and treatment of individuals with COVID-19 are more vulnerable to getting infection than the general public as well as more prone to spread infection to their loved ones and this may also results in psychological distress (22, 23). Furthermore, during the episodes of SARS and MERS multiple cases of panic attacks, anxiety, stress even suicides have been reported (3, 24–26).

The transmission of the disease among the HCP is linked with improper training, protection, not following the recommended protocols or guidelines, absence of isolation rooms and also the lack of knowledge and awareness regarding the course and spread of the disease (27). It is well documented that proper knowledge of the disease may have the positive impact on the attitude and practices thus less chances of infection (28).

Nursing is the largest healthcare profession in the world, with approximately 20 million nurses worldwide(29). Along with the other HCPs nurses plays a pivotal role in healthcare setup in prevention, infection control, isolation, continuous monitoring of the patients and because of their unique patient-facing nature, there are occupational risks to providing care during the COVID-19 outbreak (30) and it is also reported that the chances of occupational exposure is relatively higher in nurses (31). Moreover, previous studies have shown that higher level of anxiety and depression among the nurses due to their long interaction with the patients and ths nature of their job (32–34). Deprived mental health among nurses may not only be damaging to them but may also impede effect their professional routine and in turn, the quality of providing patient care (35) as it is evident that wellbeing of the mental state is crucial in order to manage infectious diseases (36, 37).

The current study aimed to assess the knowledge, practices, attitude and anxiety levels toward COVID-19 among nurses of Karachi, Pakistan who are directly involved with COVID-19 patients. The findings will help us to evaluate the knowledge and anxiety levels among the nurses associated with patients suffering from Corona Virus in different hospitals of Karachi, Pakistan and the results of this study will also help the authorities to organize the necessary educational training programs in order to provide up-to-date information and deliver the best practice to control the COVID-19 disease and to arrange counseling sessions or physiatrist/psychological intervention to control the psychological and mental problems.

## Methodology

In this study we recruited those nurses who were directly involved with patients having COVID-19 in different hospitals of Karachi, Pakistan. Due to the pandemic and lock down, an online survey on Google form was conducted with limit to single input. A validated questionnaire that was previously used in Iran (38) with few changes was sent to the eligible participants as per convenience snowball sampling. Social media platforms WhatsApp and Facebook Messenger were used to deliver the questionnaire. To every participant a time of 6 days was given from 15^th^ May 2020 to 20^th^ May 2020. Our questionnaire had questions covering demographics, knowledge, Practices and attitude. As per Lickerts scale we had questions that evaluated the nurses’ anxiety regarding their, their family infection with 2019-nCoV and anxiety performing aerosol producing procedures like taking naso pharyngeal swab, nebulizing, suctioning etc.

According to Raosoft the minimum required sample size was 67 considering a response rate of 50%, 90% confidence interval (CI) and 10% margin of error. Ethical approval was obtained from the ethics and review committee of Altamash Institute of Dental Medicine, Karachi, Pakistan (AIDM/EC/04/2020/03). The participation was voluntary, informed consent along with the approval to publish the outcomes of the study was obtained. SPSS 21 was used for data analysis. Chi Square and t-tests were applied to compare different factors. Value < 0.05 was considered significant.

## Results

Total 85 forms were submitted and due to incomplete submissions and errors we excluded 7 forms so the data of 78 nurses is analyzed.

Among the participants, 38 (48.71) were females and 40 (51.28%) were males. The difference among gender is not significant (P = 0.821) **(Table1)**

**Table 1:**
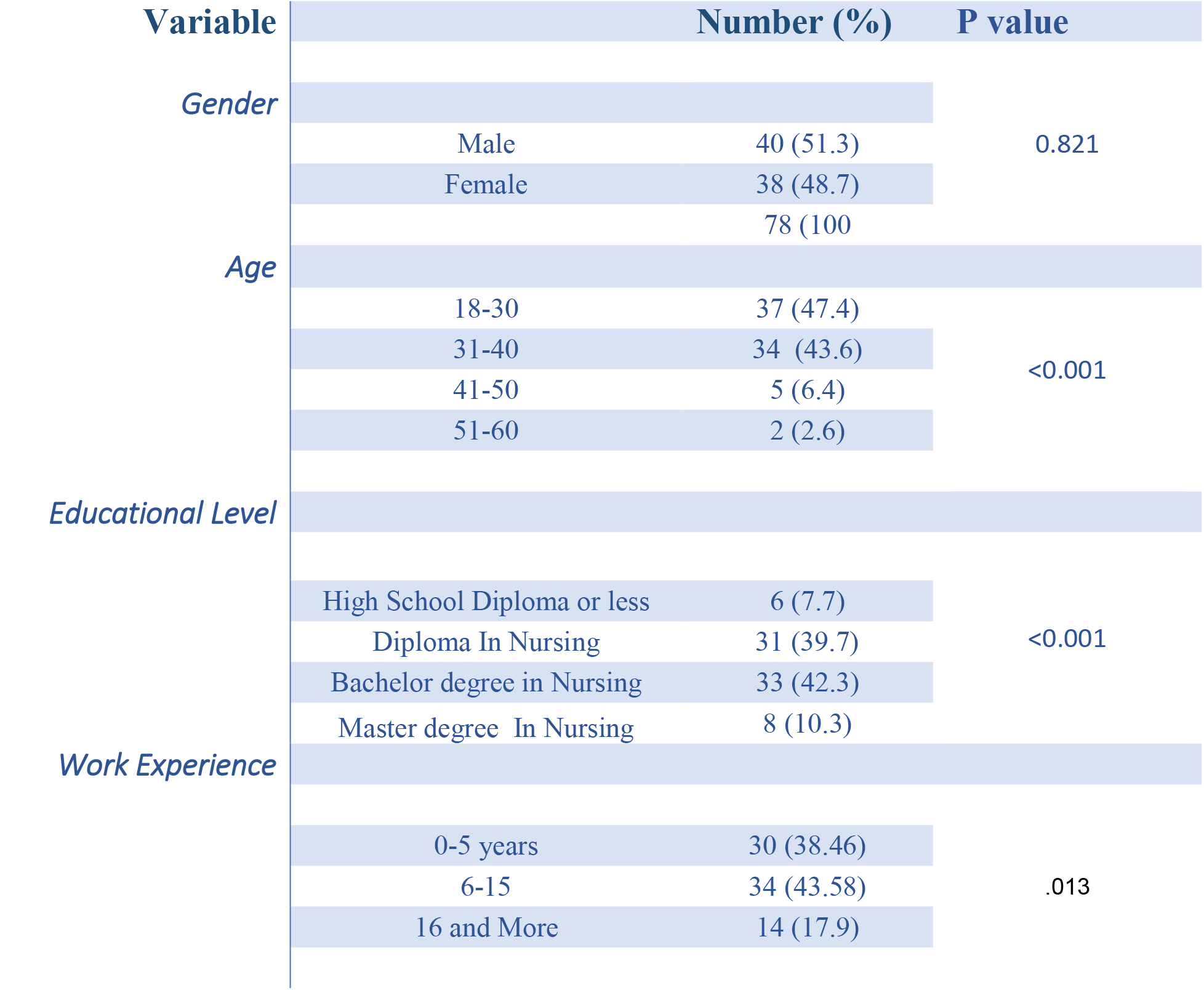
Demographic Characteristics of the Participants

In the current study 37 (47.43%) of the participants were below 30 years, 34 (43.6%) were in the age group of 31 to 40 years and 5 (6.4%) were between 41- 50years old and 2 (2.6%) participants were above 50 years. We identified that 8(10.3%) were having Master’s degree in nursing, 33 (42.3%) of the participant were holding a Bachelor‘s degree and 31 (39.7%) nurses were equipped with diploma in nursing and 6 (7.7%) were having education till high school or less. In this study 30 (38.5 %) of the nurses were having experience of less than 5 years. The majority of nurses were having experience of 5-15 years i.e. 34 (43.6%) and 14 (17.9%) were having experience of 16 years or more. There is a statistically significant difference of age, education and working experience among the participants. **(Table1)**

In the current study we explored that 12 (15.38%) of nurses reported that they were infected with COVID-19 (P>0.05) and 56(71.79%) of the participants claimed that their colleagues have been infected with the virus (P< 0.05). Family member of 14(17.94%) of the participants found positive for COVID-19 (P>0.05).

The bulk of the participants were aware about the symptoms, treatment options, routes of transmission and risk factors for CVID-19. Substantial number of the participants answered correctly **(Table 2)**. The knowledge score ranged from 5 to 19 and the mean knowledge score of nurses was 14.67 *±* 3.36 (P< 0.05). All participants knew that COVID19 is a viral disease and it is contagious. Substantial number of the participants answered correctly **(Table 2)** Three main symptoms of COVID-19 i.e. fever, shortness of breath and dry Cough were mentioned by 98.71, 98.71 and 91.02% of the nurses respectively. **(Table 2)**

**Table 2:** Knowledge of the Participants

| Questions | Options | Number (%) |
| --- | --- | --- |
| **COVID-19 Mortality Rate** | < 5% | 44 (56%.41) |
|  | 10% -20% | 19 (24.35%) |
|  | 21% - 30% | 5 (6.41%) |
|  | > 30% | 10 (12.82%) |
| **Incubation Period** | 2-14 days | 43 (55.12%) |
|  | 2-7 days | 2 (2.56%) |
|  | 7-14 days | 30 (38.56%) |
|  | 7-21 days | 3 (3.84%) |
| **Symptoms of COVID-19** | Fever | 77(98.71%) |
|  | Dry Cough | 71 (91.02%) |
|  | Runny Nose | 56 (71.79%) |
|  | Shortness of Breath | 77 (98.71%) |
|  | Joint/Muscle ache | 42 (53.84%) |
|  | Red Eyes | 35 (44.87%) |
|  | Headache | 42 (53.84%) |
|  | Diarrhea | 32 (41.02%) |
|  | Wet cough | 30 (38.46%) |
| **COVID 19 is a** | Viral Disease | 78 (100%) |
|  | Protozoal Disease | 0 (0.0%) |
|  | Bacterial Disease | 0 (0.0%) |
|  | Fungal Disease | 0 (0.0%) |
| **Route of Transmission** | Airborne | 62 (79.48%) |
|  | Direct Contact | 68 (87.17%) |
|  | Droplet | 70 (89.74%) |
|  | Indirect Contact | 56 (71.79%) |
|  | Sexual Contacts | 23 (29.48%) |
|  | Fecal-Oral Route | 29 (37.17%) |
|  | Others |  |
| **Factors considered to identify patients at risk of having COVID-19** | Presence of symptoms of Respiratory Distress | 70 (89.74%) |
|  | Presence of symptoms of Gastroenteritis | 20 (25.64%) |
|  | History of travel to areas experiencing upsurge of COVID-19 | 68 (84.61%) |
|  | History of contact with possible infected patients | 66 (84.61%) |
| **What is the routine treatment of COVID 19?** | No Treatment | 18 (23.07%) |
|  | Supportive Treatment | 66 (84.61%) |
|  | Vaccine | 6 (7.69%) |
|  | Not sure | 14 (8.97%) |
|  | Other (Onion, Steam, Ablution) | 4 ( ) |

The main sources of COVID-19 information are hospital or health ministry and social media 67(85.89%) and 47 (60.25%) respectively, followed by Colleagues OR Friends, Research Articles, Television and Radio, etc. Some of the participants were actively involved in taking webinars and reading research material on internet (**Fig 1**).

**Fig 1:**
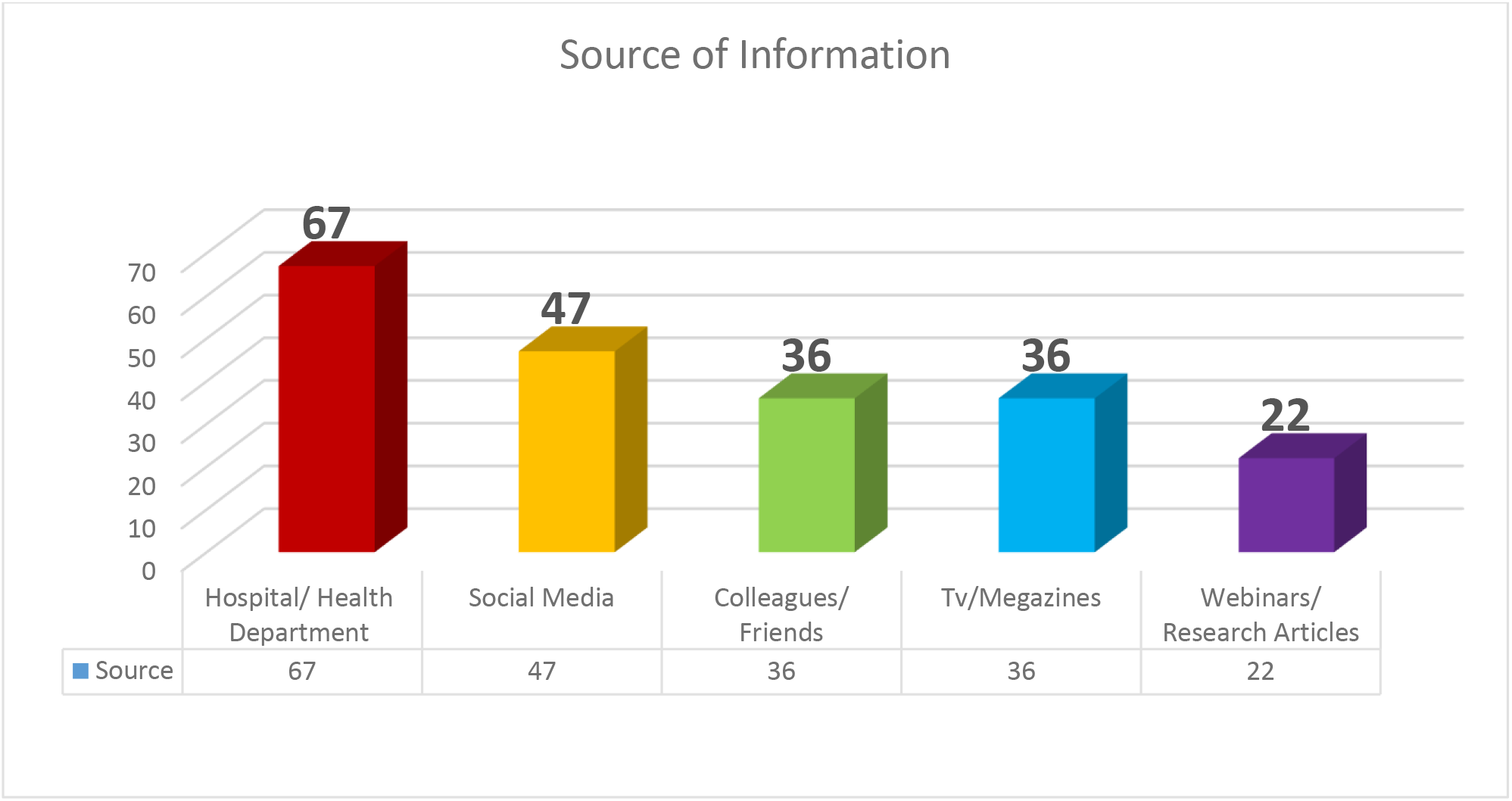
Source of Information

We analyzed that significant number of nurses have anxiety (p< 0.001). The number of nurses having no or mild anxiety are 6 (7.7%) and 6 (7.7%) respectively and those having moderate, high and very high levels of anxiety are 19 (24.4%), 24 (30.8%) and 23 (29.5%) respectively **(FIG 2)**.

**Fig 2:**
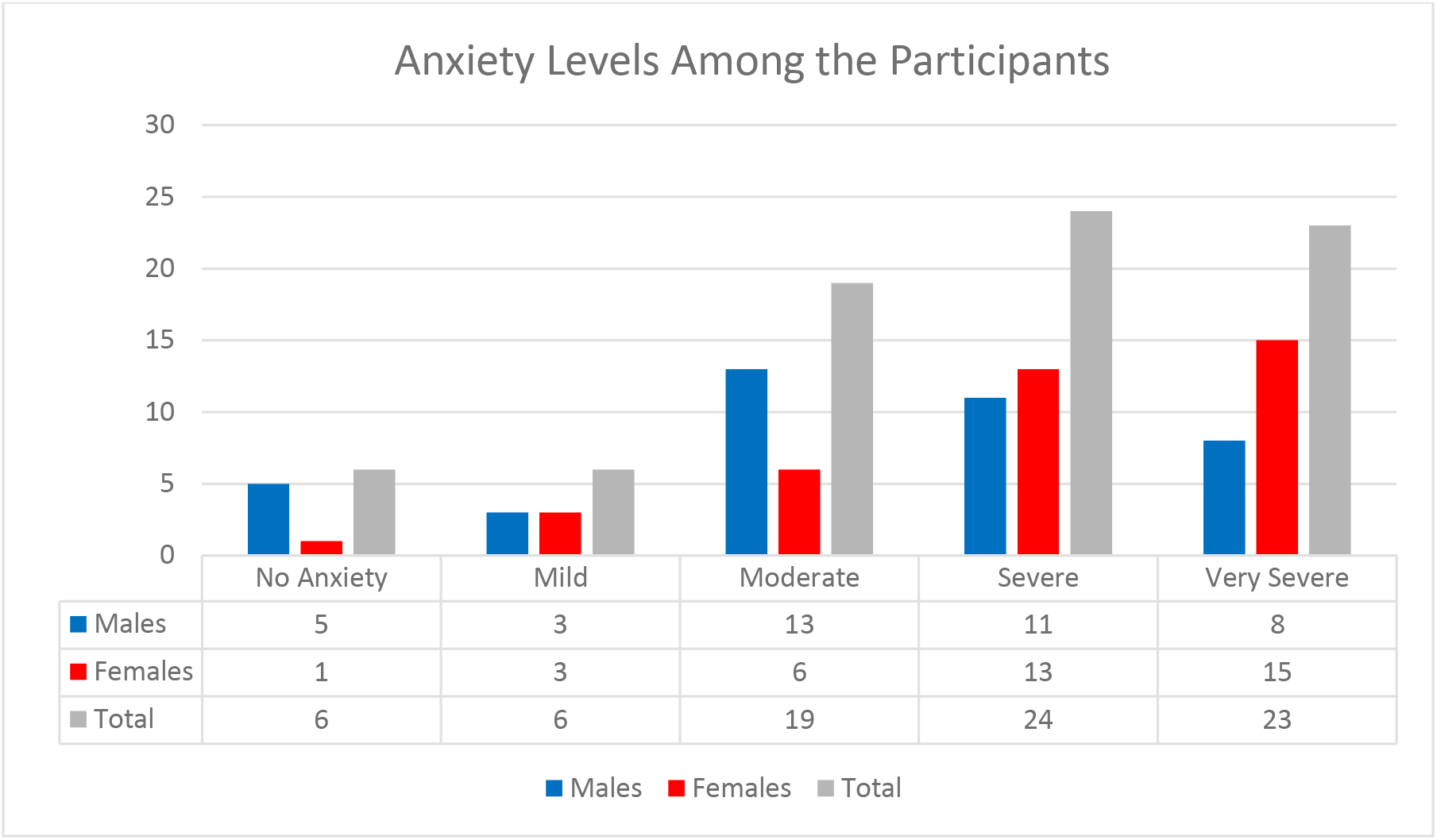
No of males, females and total number of participant with no, mild, moderate, severe and very severe anxiety.

Significant difference in the levels of anxiety was observed among the genders (p = 0.016). Females are more anxious as compared to males **(FIG 2)**. In the range of 1-5, where 1 means no anxiety and 5 means very sever anxiety, we calculated that the mean anxiety score to be infected for participant was 3.60 ± 1.25 and for their family the mean anxiety level was recorded 3.85 ± 1.31. Anxiety about performing measures like aerosol producing procedures, taking nasopharyngeal swab, nebulizing, suctioning etc. was calculated 3.30 ±1.33 **(Fig 3)**. We evaluated that the anxiety levels are significantly higher among those individuals who have been previously infected with COVID-19 (P = O.013).

**Fig 3:**
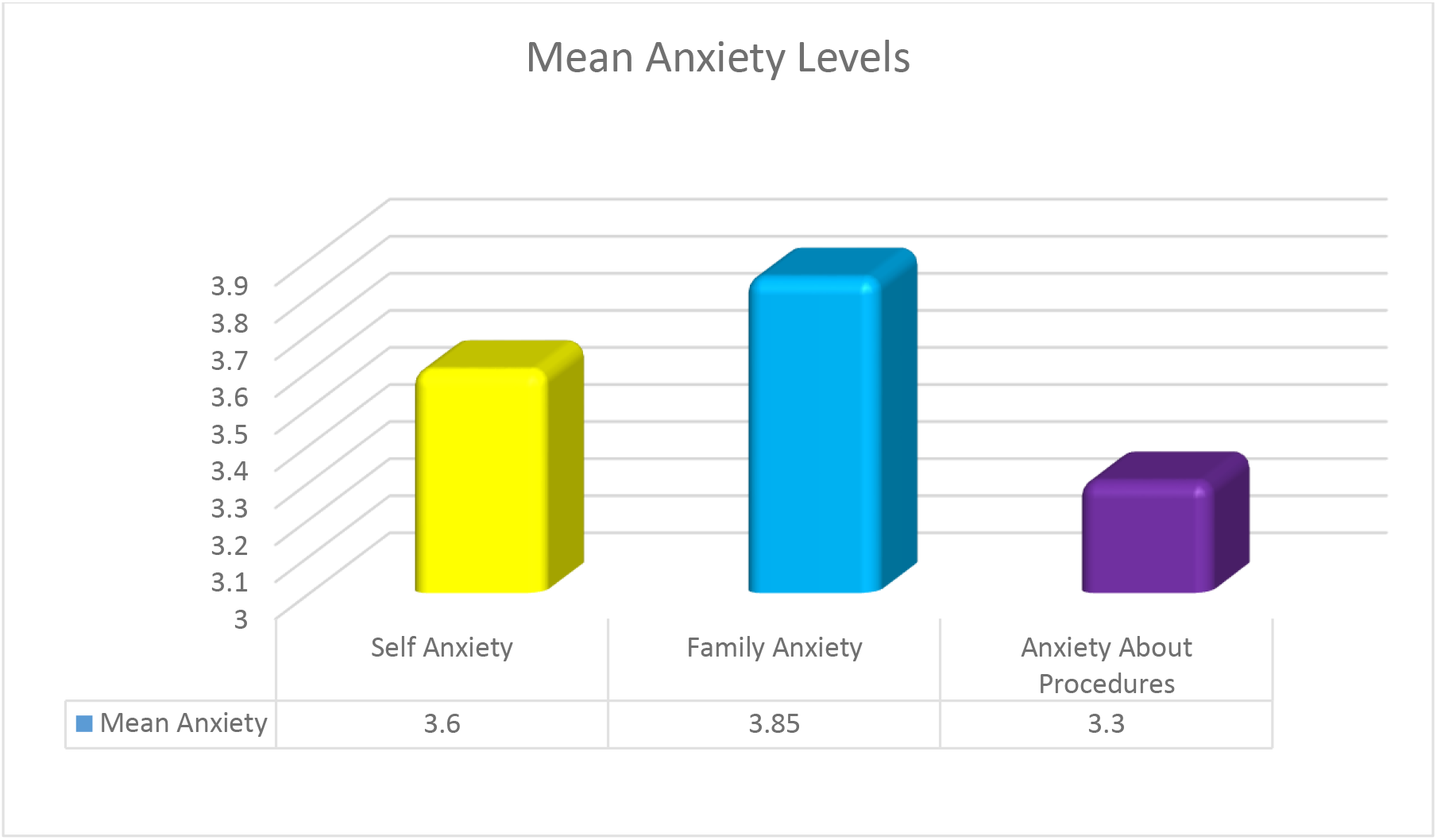
Mean Anxiety levels

As per our results, around 80% of the nurses put face masks, protective clothing, placed suspected patients in ventilated single room, routinely clean and disinfect environmental surface, frequent hand cleaning and follow respiratory hygiene and cough etiquette and practice social distancing. **(Table 3)**.

Current study findings demonstrate a positive attitude of the nurses towards COVID-19. 75(96%) nurses who have dedicated themselves for COVID-19 patients willing to continue their work. Large number of nurses 72 (92.3%) want more people to be trained to fight with COVID-19 and 59(75.6%) of the nurses rejected the idea of having attendants with the patients in the hospital **(Table 3)**.

**Table 3:** Practices and Attitude of Nurses towards COVID-19

| **Practices and Attitude of Nurses towards COVID-19** |  |  |
| --- | --- | --- |
| **Item** | **Response** |  |
|  | **Yes** | **No** |
| **Put facemask on known or suspected patients** | 75 | 3 |
| **Place known or suspected patients in adequately ventilated single rooms** | 68 | 10 |
| **All health staff members wear protective clothing** | 72 | 6 |
| **Avoid moving and transporting patients out of their area unless necessary** | 70 | 8 |
| **Routinely clean and disinfect surfaces in contact with known or suspected patients** | 67 | 11 |
| **Clean and disinfect environmental surfaces** | 64 | 14 |
| **Frequently clean hands by using alcohol-based hand rub or soap and water** | 69 | 9 |
| **Safe injection practices (i.e., aseptic technique for parenteral medications)** | 47 | 31 |
| **Respiratory hygiene / cough etiquette** | 64 | 14 |
| **Practice social distancing** | 65 | 13 |
| **Do you see the need for more people to be trained by the medical staff?** | 72 | 6 |
| **In a hospital do you prefer having more attendants with the patient?** | 18 | 59 |
| **You want to continue work with COVID 19 patients?** |  |  |
| If we get Proper Training and PPE | 42 | 3 |
| YES | 29 |  |
| If we get special allowance | 04 |  |
| Total | 75 |  |

## Discussion

To best of our knowledge, this is the first study that has targeted only the Nurses in order to assess their knowledge and practices towards COVID 19 and their level of anxiety in Pakistan. This study was being conducted when Pakistan crossed the mark of 40000 reported cases of COVID 19. In the current study, the participants were the nurses who were directly involved with COVID-19 patients. We assessed their knowledge, attitude and practice in order to shield them and prevent the further spread of the infection. It is reported that nurses are more prone to infection due to the close contact with the patients (39). Moreover, we analyzed the anxiety levels among the nurses as studies suggests that in case of big disasters mental health issues may arise (40).

In this study the data of 78 participants was analyzed. Total number of males were 40 and females were 38. There is no significant difference among the gender. Usually in other studiers higher female to male ratio has been observed(31) but contrary to other studies in our study we found higher number of males. This is because of the reason that in Pakistan males are usually the bread earners and they have no other option on the other hand many female nurses have quit their jobs and resigned due to the family burden(41).

In this study we observed a significant difference in age, educational level and experience. We further investigated that 90% of the nurses are below 40 years of age and majority have bachelor’s degrees or associated diplomas in nursing. Similar finding witnessed in other studies (38, 42).

The finding of the current study showed that nurses in Karachi have sufficient knowledge regarding different aspects of COVID-19. In a recent study conducted on health care professionals in Pakistan reveled that HCP have good knowledge towards COVID-19 (43, 44). Moreover different studies performed in different parts of the world showed the strikingly similar results (38, 45, 46). Contradictory to the findings of our study, a research conducted in Pakistan few weeks before reported low level of knowledge among the nurses(47). Previous studies in different parts of the world during epidemic revealed that the rate of influx of information is much high during the current pandemic(48, 49).

We appraised that Health department or hospital and social media and are the two major sources of information regarding COVID 19. Our results are further endorsed by different studies stated that the previously mentioned sources are the primary sources of information regarding COVID-19 (42, 44, 50).

In the current study we have observed that nurses’ followed correct protocols in order to prevent themselves and patients. Such practices are very important in order to control the cross infection. Practices are associated with knowledge, work experience, and other factors(51).

In this study, 57 (73.07%) participants have received formal training to cater the COVID-19 positive patients and infection control. Contrary to our findings the previous study conducted in Pakistan showed that 50% of the nurses had not undergone any training and have limited knowledge about COVID-19(47). It is also possible that the training could be the reason of increased awareness and knowledge among nurses which is evident in the current study. Zhong et al stated that with good knowledge and practice and following the protective measures, HCPs not only save themselves but show good message of awarness to the community (51).

We investigated that 12 (15.38%) nurses were infected with COVID 19 and 56(71.79%) of the nurses have colleague(s) that had been infected with the COVID 19. Moreover 14(17.94%) nurses reported that their family member(s) had been infected. HCPs show high prevalence of getting infection with any contagious or non-contagious infection (52). Furthermore data from other countries are in accordance with our findings(53) In Italy 12 % of the nurses got COVID-19. In

Brazil it is also reported that 1000s of nurses have been infected and 100s dead (54). Same are the conditions in other countries as per reports (55–57) Moreover we believe that higher rate of infection and mortality among the healthcare professional and their family members could be one of the cause of anxiety. But contrary to our findings, a study in Iran showed totally different results with zero infected case among nurses (38).

Likewise in different researches, in our study significant majority of the participants believes that there is dire need of more trained workers in order to accommodate the need that is aroused because of the current pandemic (58, 59).

Anxiety is an undesirable emotional condition perceived individually and it is reported that anxiety is one the commonest psychological hurdle of nurses (31). In our study 72 (91.3%) nurses have the overall anxiety ranges from mild to very severe. In a similar study on health care workers conducted in China expressed similar results (60) and in another study that was purely conducted over nurses showed a higher level of anxiety among the nurses which further validates our findings, furthermore in accordance with the results of our study it also showed the anxiety for the family member (38, 61, 62). Moreover previous studies have also shown higher level of anxiety and depression among the nurses due to their long interaction with the patients and nature of their job (32).Similar findings were reported by other studies during SARS and MERS epidemics (3, 63, 64). According to the results of this study females have more anxiety as compare to males which is in agreement to the outcome of other studies (65, 66).

In our opinion the high rate of anxiety among the nurses is because of unavailability of personal protective equipment (PPE), lack of counselling sessions, deficiency of proper health care facilities etc. Moreover job stress, demands, exertions may not only have negative impact on the psychology and mental health but also on the general wellbeing(67, 68). Moreover due to the social media and easy access to the current news and information that the anxiety and obsession has been increased (69, 70). Anxiety of nurses towards their family member to be infected with COVID-19 is significantly on the very severe side. Studies have shown that due to the highly contagious nature of this virus healthcare professionals are in stress and depression as they might transfer the infection to their family members(45). In order to reduce the anxiety counseling sessions should be organized. Same recommendations were given in parallel studies as the wellbeing of the mental state is crucial to in order to manage infectious diseases (36, 37).

## Conclusion

This study concludes that the Nurses working with COVID-19 patients have good knowledge regarding symptoms, route of infection, treatment options etc. Majority of the Nurses in Karachi, Pakistan showed typical signs of anxiety. Therefore it is recommended that proper counseling sessions may help them cope with the pandemic. Proper training and mentorship along with fortnightly or monthly refresher course may aid into tackling such situations.

## Data Availability

We will made all the data of this manuscript available.

## Conflict of Interest

The authors declare that they do not have any interests that could constitute a real, potential or apparent conflict of interest with respect to their involvement in the publication

## Funding

None

## Acknowledgements

We acknowledge the support of the ethic and review committee of Altamash Institute of Dental Medicine, Karachi, Pakistan. We would like to extend heartfelt graciousness to Dr. Shoaib Durrani from Durrani Dental Clinic, Karachi, Pakistan, Mr. Muhammad Saqlain of Quaid –e-Azam University, Pakistan and MS. Marzieh Nemati, Bahareh Ebrahimi and Fatemeh Nemati of Shiraz University of Medical Sciences, Iran and to all the participants and people who provided support at every step of the research.

## Strength and Limitations

This study emphasized on the less discovered field where very limited literature was available. The sample size was limited as the study only comprises of the nurses from Karachi, Pakistan working with COVID-19 so the results cannot be generalized. It was an online survey so factors like dis honest answers and biasness should be considered.

## Author’s contribution

AS and MM conceived the study and designed the questionnaire. HM, SM and SH collected data. SM and SH performed statistical analyses and prepared tables and figures. MM, FH and AS drafted the manuscript. AS reviewed the manuscripts. MM supervised the project and is responsible for the integrity of the research. All authors contributed comprehensively contributed in writing and critically revised and approved the final draft of the manuscript.

## Notes

### Competing Interest Statement

The authors have declared no competing interest.

### Author Declarations

Ethical approval was obtained from the ethics and review committee of Altamash Institute of Dental Medicine, Karachi, Pakistan (AIDM/EC/04/2020/03).

